# Physical therapists’ perceptions of telerehabilitation for patients with musculoskeletal conditions in a post-pandemic world

**DOI:** 10.1101/2025.01.17.25320739

**Authors:** Kevin McLaughlin, Kate I. Minick, Julie M. Fritz, Nicole Tannahill, Anne Spar, Marissa Feinsilver, Matthew Weber, Erika Opoku, Jeff Adams, Richard L. Skolasky

## Abstract

**Objective:** To examine physical therapists’ experience providing telerehabilitation and their perceptions of telerehabilitation for patients with musculoskeletal conditions.

**Methods:** Survey of members of the Academy of Orthopaedic Physical Therapy

**Results:** We received 208 completed responses to our survey invitation. Most physical therapists responding to our survey reported using little to no telerehabilitation prior to the pandemic but increased use during the pandemic, followed by decreased use of telerehabilitation after the pandemic. Respondents reported using telerehabilitation slightly more after the pandemic than beforehand. Physical therapists reported that they believed they could deliver effective care using telerehabilitation but that it was not as effective as in-clinic care and they would prefer to use telerehabilitation as part of a hybrid care model versus using only telerehabilitation. Physical therapists suggested that certain patients were more likely than others to benefit from telerehabilitation and identified specific factors (e.g., patient preference, self-efficacy, postoperative status) that influenced whom they would consider to be most appropriate for telerehabilitation. “Patient issues with technology” was identified as the most common barrier to telerehabilitation use. Most physical therapists reported that they planned to use telerehabilitation in the future and agreed that telerehabilitation plays an important role in expanding access to physical therapy in the United States.

**Conclusions:** Physical therapists believed that telerehabilitation continues to play an important role in a post-pandemic world. Although physical therapists reported that they considered telerehabilitation to be an effective method for delivering care, they did not consider it to be a replacement for in-clinic care and believed that certain patients are more likely than others to benefit from this approach.

**Impact Statement:** Physical therapists consider telerehabilitation to be a viable care option for patients with musculoskeletal pain.

## Introduction

Prior to the COVID-19 pandemic, barriers that prevented the widespread use of telerehabilitation included state practices acts that prohibited physical therapists from delivering care via telerehabilitation and payer policies that did not allow for reimbursement for telerehabilitation services provided by physical therapists.^1,2^ However, policy changes during the early stages of the pandemic facilitated rapid expansion of telerehabilitation,^3^ with reports indicating that as many as 12% of physical therapy patients accessed telerehabilitation at the peak of its use in 2020.^4^ A number of studies were also conducted during that time that found telerehabilitation to be acceptable and effective among patients with a variety of health conditions, including those with musculoskeletal pain.^5–9^

Although telerehabilitation played a clear role in expanding access to physical therapy during the pandemic, the role of telerehabilitation in a post-pandemic world is less clear. Since its peak in the first quarter of 2020, telerehabilitation utilization rates have decreased substantially.^4^ Multiple factors may have influenced this decline, such as changes in social-distancing guidelines, patient demand for these services, or uncertainty surrounding reimbursement for telerehabilitation in the future. Studies conducted during the pandemic reported that physical therapist perceptions of telerehabilitation served as both enablers of and barriers to the adoption of telerehabilitation into practice.^10,11^ It stands to reason that physical therapists’ perceptions of telerehabilitation will continue to play a major role in the way that telerehabilitation is used after the pandemic. However, there has been little to no research examining physical therapist perceptions of telerehabilitation since the end of the public health emergency.

To address this knowledge gap, we conducted a national survey examining physical therapists’ perceptions of telerehabilitation for patients with musculoskeletal pain. Musculoskeletal conditions are among the most common conditions seen by physical therapists, and patients with these conditions represented the largest group of telerehabilitation users during the pandemic.^4,12,13^

## Methods

### Survey Development

A survey was developed for the purposes of this study by members of the study team with expertise in physical therapy and musculoskeletal health and experience with telerehabilitation. After development, the survey was piloted among 6 physical therapists with experience providing care to patients with musculoskeletal pain and telerehabilitation. The final version of the survey incorporated feedback from the pilot regarding survey length, content, and clarity.

The final version of the survey used in this study consisted of 29 questions, separated into 5 sections: (1) current and previous use of telerehabilitation, (2) perceptions of telerehabilitation, (3) future use of telerehabilitation, (4) patient subgroups appropriate for telerehabilitation, and (5) respondent characteristics. Questions included a combination of multiple choice and Likert-style response options. One question included an “other” response with a free-text option. The final version of the survey is available in **Supplemental Material**.

### Survey Distribution

Surveys were mailed electronically to members of the Academy of Orthopaedic Physical Therapy (AOPT). This provider population was chosen as it comprises primarily physical therapists with an interest in musculoskeletal conditions, our area of interest for this study. The AOPT mailing list is commonly used to conduct studies of this nature.^14–16^ Surveys were distributed in March 2024. We accepted survey responses completed by the end of May 2024.

### Data Analysis

Descriptive statistics were used to describe responses to all included survey questions. Responses to questions about respondents’ perceptions of telerehabilitation (questions 7–13), which included 5 Likert-scale response options (strongly agree, agree, neutral, disagree, strongly disagree), were collapsed into 3 categories (agree, neutral, disagree) to simplify comparisons between questions. Only complete survey responses were included in our analysis, defined by a respondent navigating through all questions and clicking “survey complete.”

### Ethics Approval

This study was recognized as exempt by the Johns Hopkins Medicine Institutional Review Boards (IRB00402769).

## Results

### Respondent Characteristics

We received a total of 218 responses to our survey invitation, with 208 physical therapists responding to all survey questions (95.4%). The sample of physical therapists who completed our survey were experienced, with over half of respondents (n = 107; 51.7%) reporting over 20 years of experience as physical therapists and 54 (26.1%) respondents reporting between 10 and 20 years of experience (Table 1). Over half of respondents were board-certified specialists (n = 132; 64.1%), with orthopaedics being the most common type of board certification (n = 126, 96.9%). Respondents practiced in diverse geographic settings with 104 (50.7%) respondents reporting they practiced in suburban areas, 58 (28.3%) in urban areas, and 43 (21.0%) in rural areas.

**Table 1.**
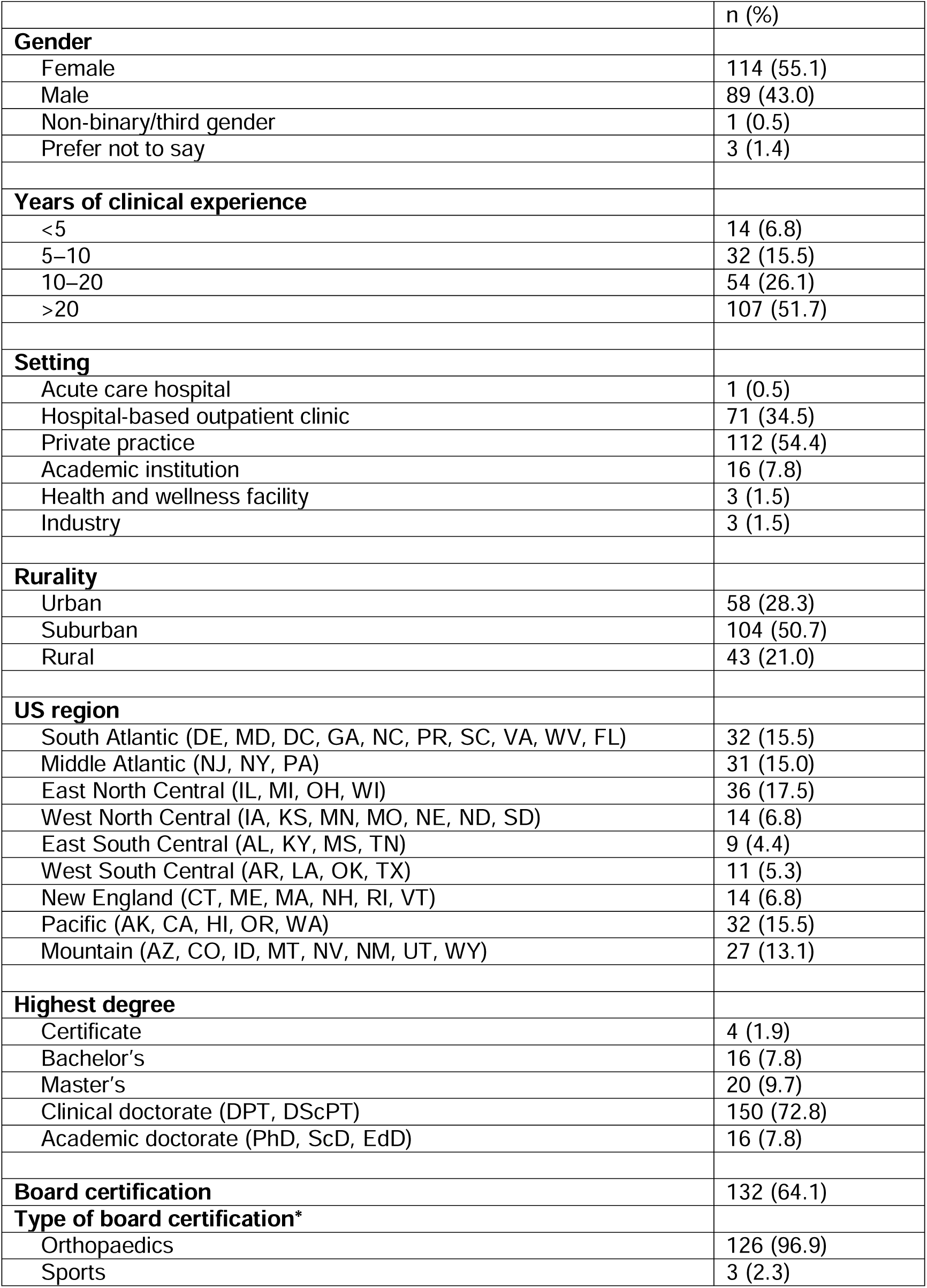
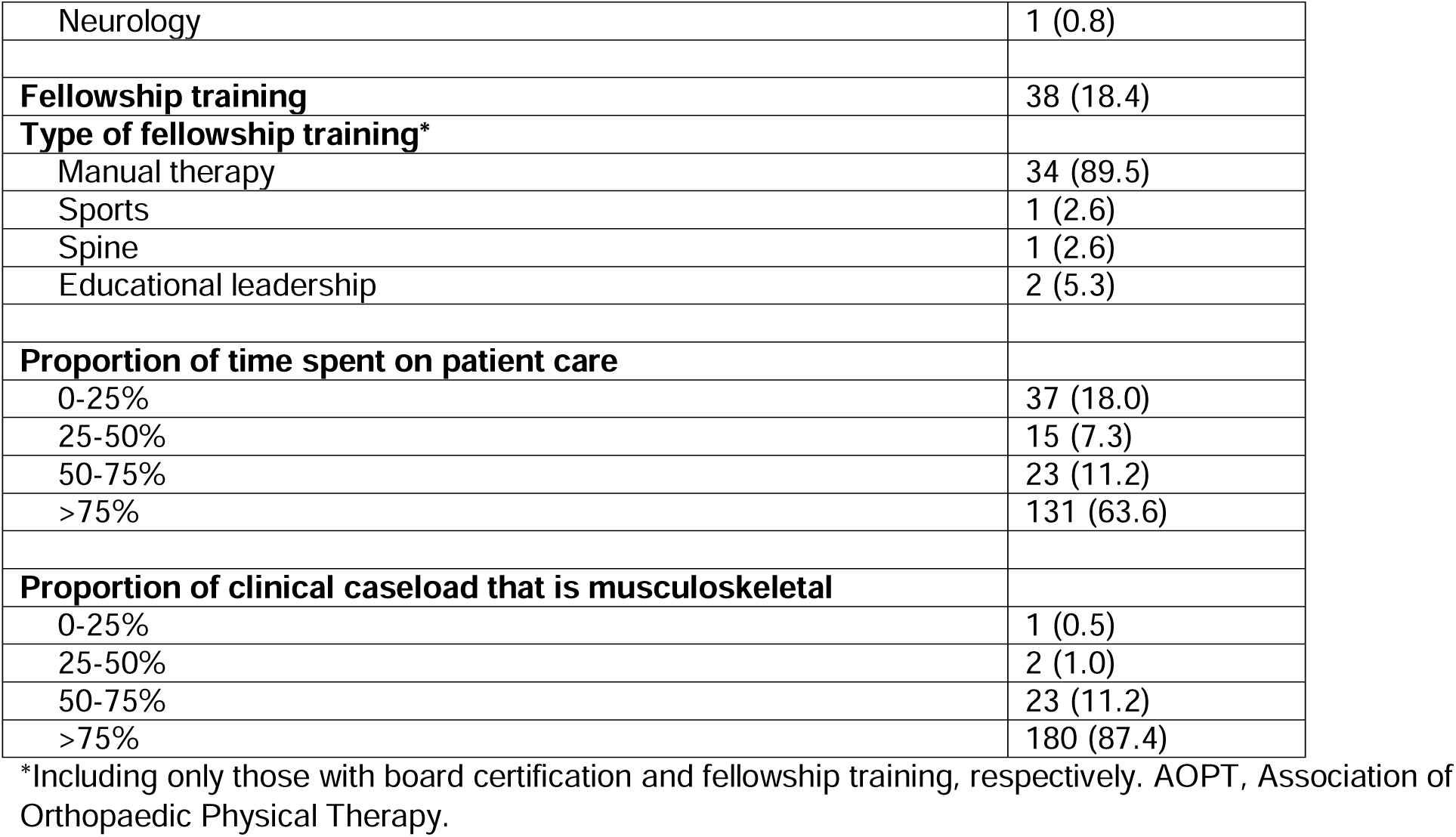
Characteristics of AOPT Member Respondents To Telehealth Utilization Questionnaire (N = 208)

|  | n (%) |
| --- | --- |
| **Gender** |  |
| Female | 114 (55.1) |
| Male | 89 (43.0) |
| Non-binary/third gender | 1 (0.5) |
| Prefer not to say | 3 (1.4) |
| **Years of clinical experience** |  |
| <5 | 14 (6.8) |
| 5–10 | 32 (15.5) |
| 10–20 | 54 (26.1) |
| >20 | 107 (51.7) |
| **Setting** |  |
| Acute care hospital | 1 (0.5) |
| Hospital-based outpatient clinic | 71 (34.5) |
| Private practice | 112 (54.4) |
| Academic institution | 16 (7.8) |
| Health and wellness facility | 3 (1.5) |
| Industry | 3 (1.5) |
| **Rurality** |  |
| Urban | 58 (28.3) |
| Suburban | 104 (50.7) |
| Rural | 43 (21.0) |
| **US region** |  |
| South Atlantic (DE, MD, DC, GA, NC, PR, SC, VA, WV, FL) | 32 (15.5) |
| Middle Atlantic (NJ, NY, PA) | 31 (15.0) |
| East North Central (IL, MI, OH, WI) | 36 (17.5) |
| West North Central (IA, KS, MN, MO, NE, ND, SD) | 14 (6.8) |
| East South Central (AL, KY, MS, TN) | 9 (4.4) |
| West South Central (AR, LA, OK, TX) | 11 (5.3) |
| New England (CT, ME, MA, NH, RI, VT) | 14 (6.8) |
| Pacific (AK, CA, HI, OR, WA) | 32 (15.5) |
| Mountain (AZ, CO, ID, MT, NV, NM, UT, WY) | 27 (13.1) |
| **Highest degree** |  |
| Certificate | 4 (1.9) |
| Bachelor's | 16 (7.8) |
| Master's | 20 (9.7) |
| Clinical doctorate (DPT, DScPT) | 150 (72.8) |
| Academic doctorate (PhD, ScD, EdD) | 16 (7.8) |
| **Board certification** | 132 (64.1) |
| **Type of board certification\*** |  |
| Orthopaedics | 126 (96.9) |
| Sports | 3 (2.3) |
| Neurology | 1 (0.8) |
| **Fellowship training** | **38 (18.4)** |
| **Type of fellowship training\*** |  |
| Manual therapy | 34 (89.5) |
| Sports | 1 (2.6) |
| Spine | 1 (2.6) |
| Educational leadership | 2 (5.3) |
| **Proportion of time spent on patient care** |  |
| 0-25% | 37 (18.0) |
| 25-50% | 15 (7.3) |
| 50-75% | 23 (11.2) |
| >75% | 131 (63.6) |
| **Proportion of clinical caseload that is musculoskeletal** |  |
| 0-25% | 1 (0.5) |
| 25-50% | 2 (1.0) |
| 50-75% | 23 (11.2) |
| >75% | 180 (87.4) |

\*Including only those with board certification and fellowship training, respectively. AOPT, Association of Orthopaedic Physical Therapy.

### Previous Use of Telerehabilitation

Most respondents (n = 181; 87.0%) reported that they had never provided care using telerehabilitation prior to the pandemic, with the next most common response being that respondents provided care using telerehabilitation “rarely” before the pandemic, operationalized as less than monthly (Figure 1). Most respondents indicated that, during that timeframe, patients receiving telerehabilitation care accounted for 0% of their weekly caseload (n = 182; 87.9%), followed by 1–10% of their weekly caseload (n = 19; 9.2%) (Figure 2).

**Figure 1.**
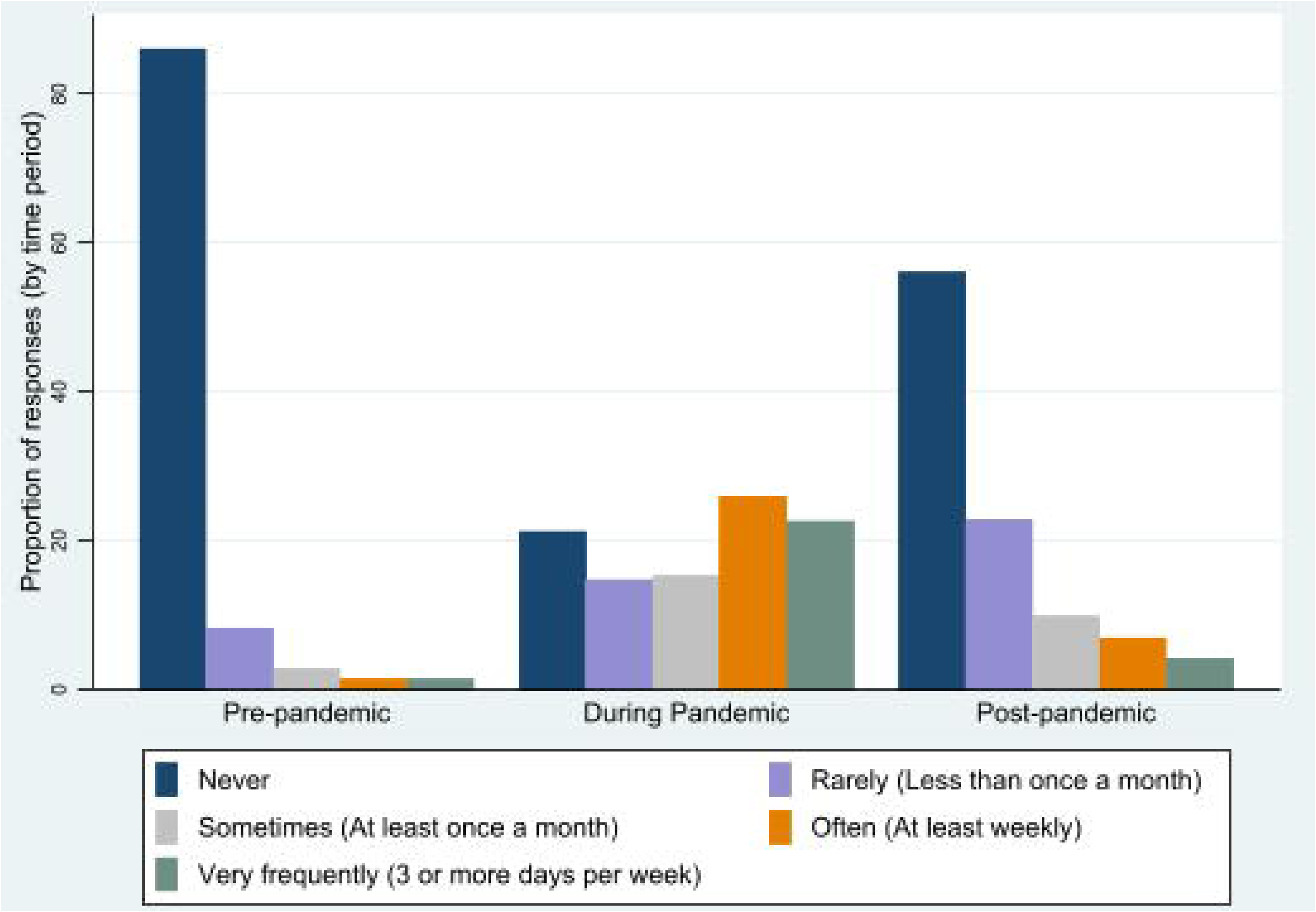
Frequency of telerehabilitation delivery before, during, and after the COVID-19 pandemic. Participants were asked to indicate the highest frequency during the period defined as March 2020–May 2023.

**Figure 2.**
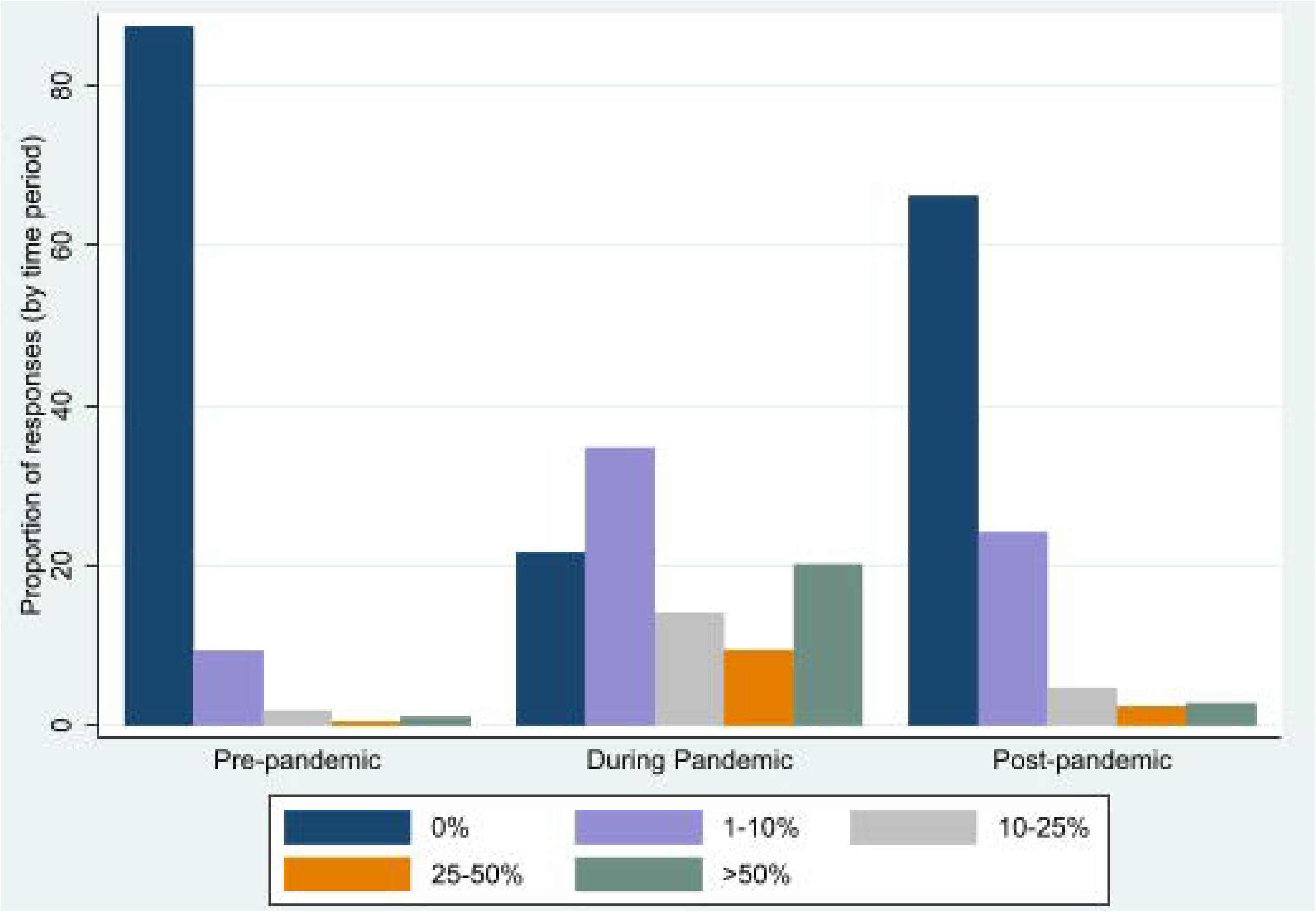
Proportion of patients utilizing telerehabilitation before, during, and after the COVID-19 pandemic. Participants were asked to indicate the highest proportion during the period defined as March 2020–May 2023.

When asked how frequently they provided telerehabilitation during the pandemic, the highest proportion of respondents (n = 55; 26.4%) indicated that they provided care using telerehabilitation “often,” operationalized in our survey as ≥1 time per week (Figure 1). The second most common response (n = 47; 22.6%) to this question was “very frequently,” operationalized as providing care using telerehabilitation ≥3 days per week. When asked what proportion of patients they saw via telerehabilitation during the pandemic, the highest proportion of respondents indicated that they provided care via telerehabilitation for 1–10% of their caseload (n = 73; 35.6%), followed by 0% (n = 44; 21.5%), and >50% (n = 41; 20.0%) (Figure 2).

When asked how frequently respondents provide care using telerehabilitation after the pandemic, the most common response (n = 117; 56.8%) was “never,” followed by “rarely” (n = 47; 22.8%) (Figure 1). Most respondents indicated that the proportion of their patients who used telerehabilitation on a weekly basis was 0% (n = 139; 66.8%), followed by 1–10% (n = 49; 23.6%) (Figure 2).

### Perceptions of Telerehabilitation

Participant responses to questions about their overall perceptions of telerehabilitation are summarized in Figure 3. The highest proportion of respondents (n = 101; 48.6%) indicated that they believed they could deliver effective care using telerehabilitation, but a majority of respondents (n = 129; 62.0%) indicated that they did not consider telerehabilitation to be as effective as in-clinic physical therapy. Most physical therapists (n = 96; 46.4%) agreed that some patients may respond better to telerehabilitation than to in-clinic care.

**Figure 3.**
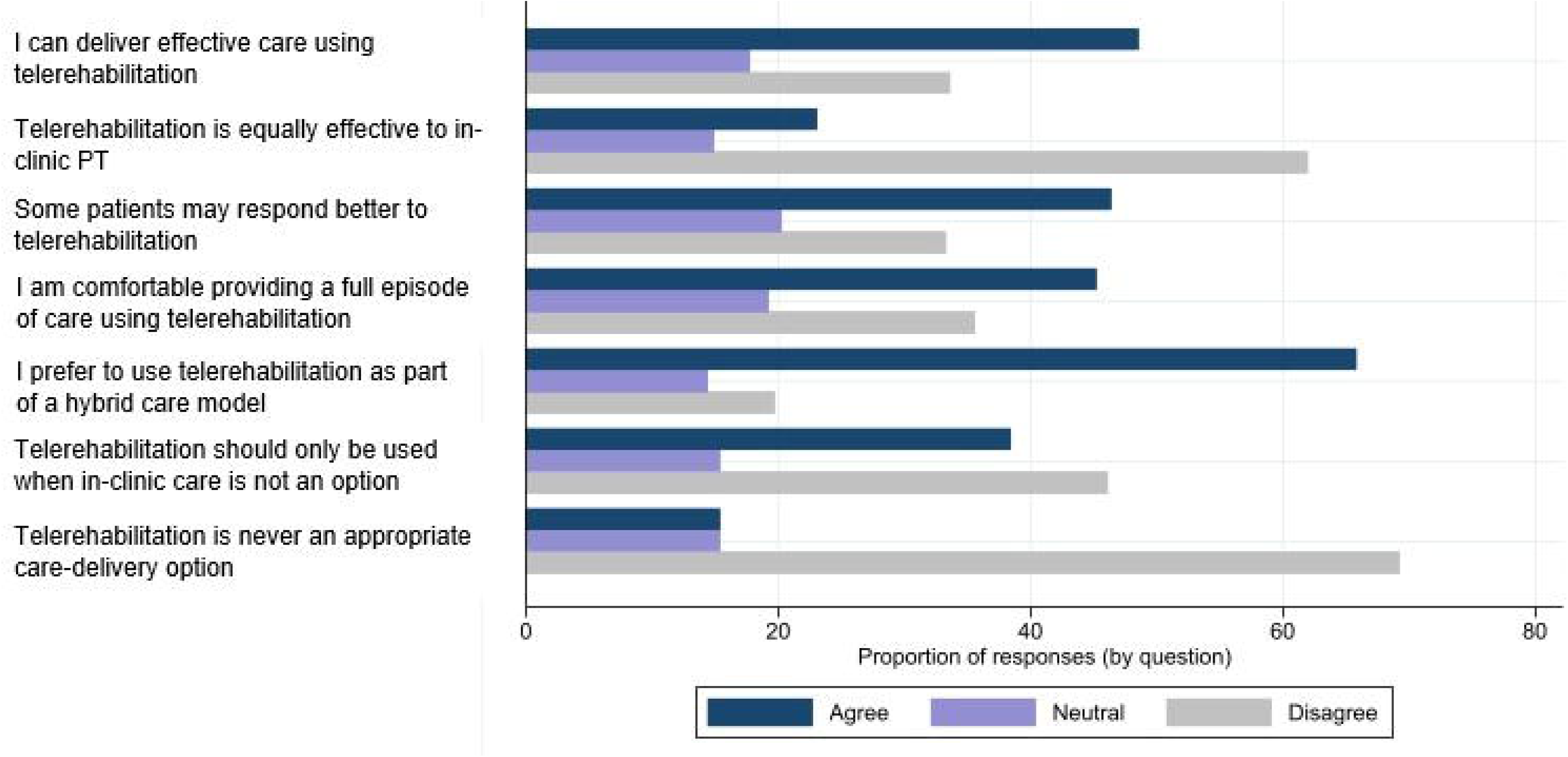
Physical therapist perceptions of telerehabilitation effectiveness and appropriateness.

With regard to the way that telerehabilitation is used as part of a full plan of care, 94 (45.2%) respondents indicated that they would feel comfortable providing a full episode of care entirely through telerehabilitation. At the same time, 137 (65.9%) respondents indicated that they would prefer to provide telerehabilitation as part of a hybrid care model if this were an option.

Most respondents (n = 96; 46.2%) disagreed with the statement that telerehabilitation should be used only when in-clinic care is not an option. Most (n = 144; 69.2%) also disagreed with the statement that telerehabilitation was never an appropriate way to deliver care.

### Future Use of Telerehabilitation

When asked about future use of telerehabilitation, the highest proportion of respondents (n = 46; 22.3%) indicated that they “agree” with a statement suggesting they plan to provide telerehabilitation in the future (Table 2). The most common response (n = 84; 40.4%) was to “agree” with a statement suggesting that telerehabilitation plays an important role in expanding access to physical therapy.

**Table 2.**
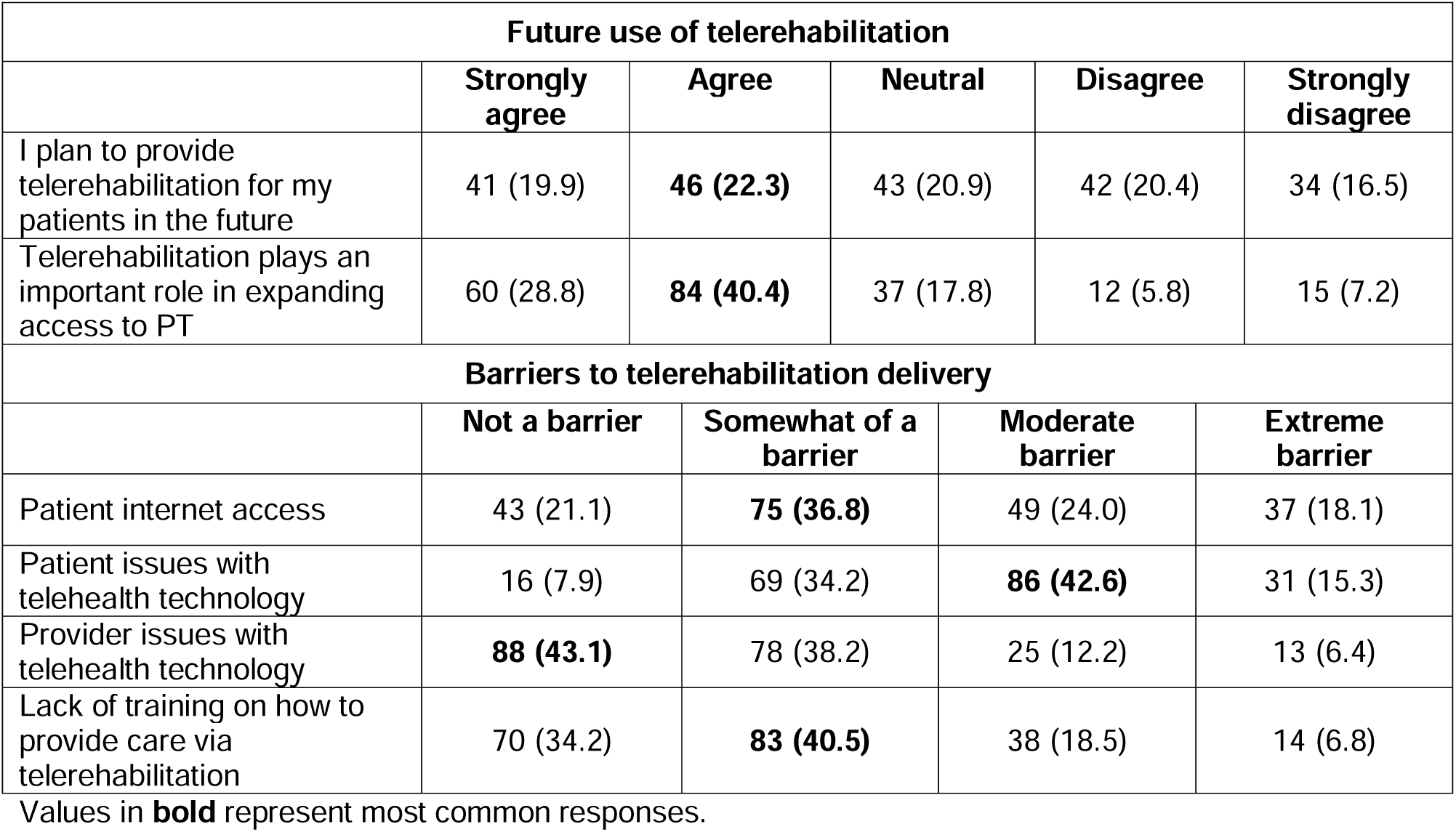
Future use of telerehabilitation and potential barriers to use.

Respondents indicated that “patient issues with technology” was a barrier to providing telerehabilitation, with 86 (42.6%) respondents indicating this was a “moderate barrier” (Table 2). Respondents indicated that internet access, provider issues with technology, and lack of provider training were less likely to be barriers, with respondents indicating these were only “somewhat of a barrier” or “not a barrier.”

### Individual Patient Factors Influencing Appropriateness for Telerehabilitation

In response to a statement that telerehabilitation is best suited for specific groups of patients with musculoskeletal pain, the most common response (n = 85; 40.9%) was “agree.” According to survey responses, providers believed patients who prefer video visits (n = 165, 80.1%), have access to the internet (n = 98, 47.8%), have high self-efficacy (n = 123, 60.3%), and are comfortable using technology (n = 145, 70.7%) are more appropriate for telerehabilitation (Table 3). Respondents indicated that they believed patients who are available for in-clinic appointments (n = 127, 62.0%), have acute conditions (n = 112, 54.9%), have high psychosocial risk (n = 77, 38.1%), have low self-efficacy (n = 122, 59.8%), or are postoperative (n = 144, 70.2%) may be less appropriate for telerehabilitation. Respondents indicated that other individual factors included in our survey had no impact on patients’ appropriateness for telerehabilitation.

**Table 3.**
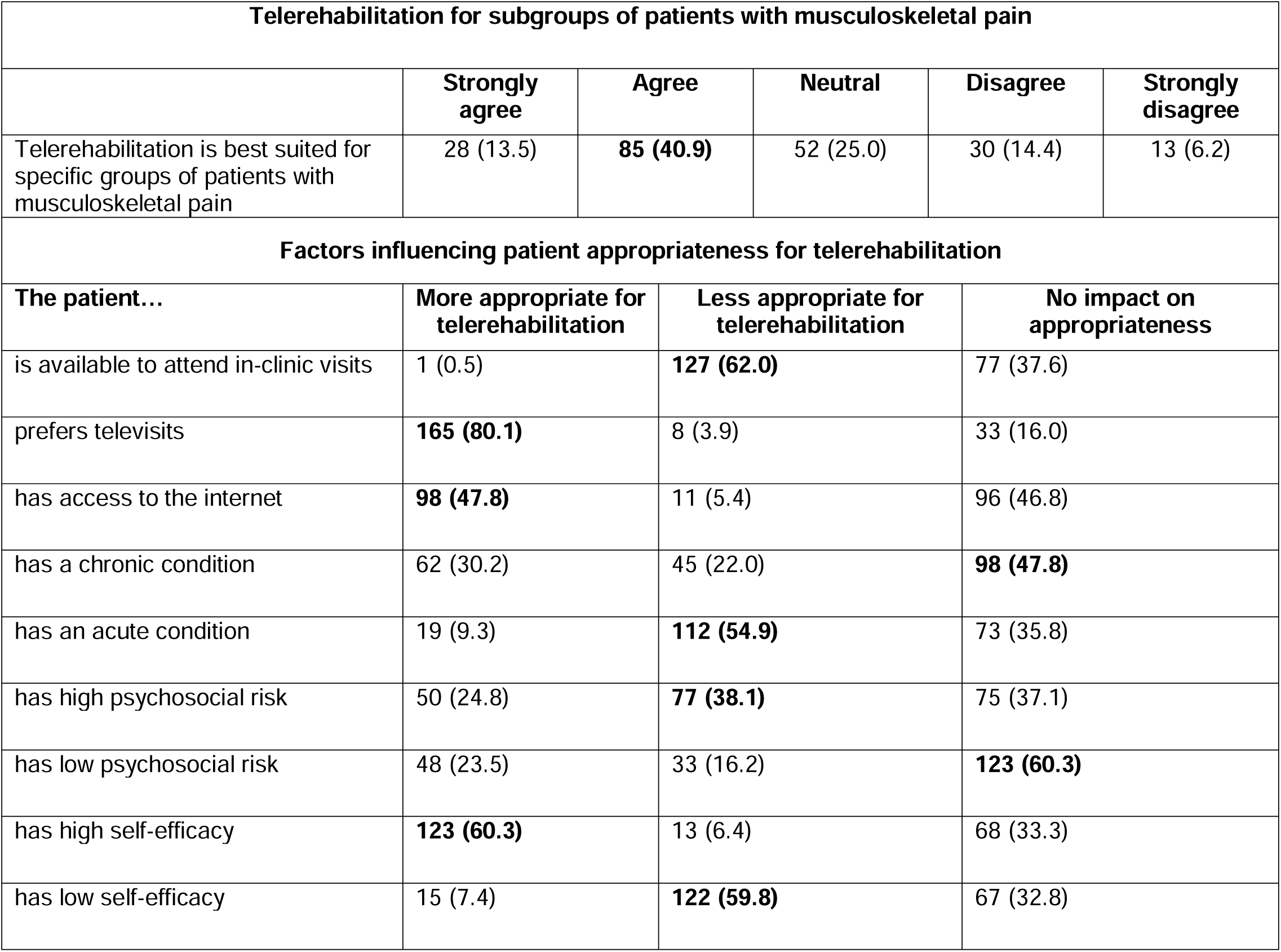
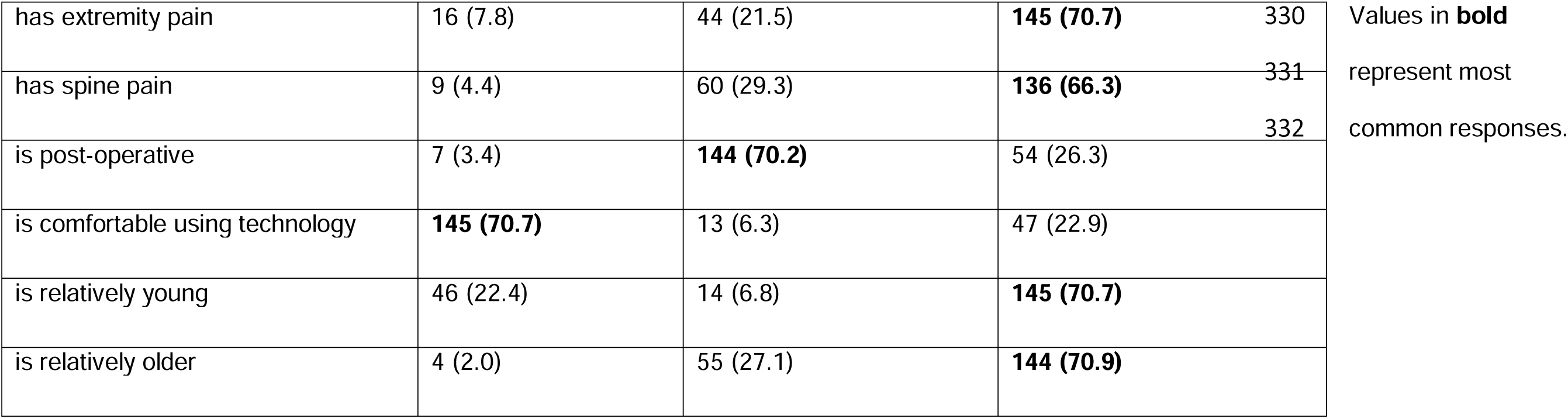
Individual factors influencing patient appropriateness for telerehabilitation.

We also provided a free-text option for respondents to indicate other factors they believed might influence patients’ appropriateness for telerehabilitation. Common factors listed by respondents included social determinants of health, safety (e.g., fall risk), medical complexity, access to technology, caregiver support, and likelihood that patients would benefit from hands-on interventions.

## Discussion

This study is one of the first to examine physical therapist perceptions of telerehabilitation since the end of the COVID-19 pandemic. The results of this study indicated that physical therapists felt they can deliver effective care for patients with musculoskeletal pain through telerehabilitation. However, most did not believe that care provided through telerehabilitation is as effective as care provided in clinic. Our results also suggest that physical therapists considered the method by which patients receive care (i.e., in-person or using telerehabilitation) to be a multifactorial decision that includes consideration of patient preference and individual patient characteristics.

The results of this study are consistent with those of previous studies that have reported an increase in the use of telerehabilitation during the early phases of the pandemic and a decline in telerehabilitation utilization during later stages of the pandemic, based on data from insurance claims and a physical therapy outcomes database.^4,17^ Our results build on these previous studies by examining telerehabilitation use after the end of the public health emergency, providing insights into its use following the pandemic. It is not surprising that our respondents reported they provide care through telerehabilitation less often since the pandemic ended. However, survey results suggested that telerehabilitation utilization rates have not returned to pre-pandemic rates. When surveyed on pre-pandemic use of telerehabilitation, 87.0% of physical therapists reported that they “never” provide telerehabilitation. When surveyed on post-pandemic telerehabilitation use, 56.8% of therapists reported that they “never” provide telerehabilitation. This suggests that approximately 30% more of the physical therapists surveyed are providing telerehabilitation after the pandemic than before the pandemic.

The results of the present study suggest that physical therapists view telerehabilitation as a useful tool for patients with musculoskeletal pain but are skeptical about its use as a replacement for in-clinic care. Multiple factors may contribute to this viewpoint; for example, physical therapists may perceive a lack of hands-on interventions (e.g., manual therapy, modalities) as a limitation to telerehabilitation, which is supported by the comments provided by physical therapists indicating that patients who stand to benefit from hands-on care are less appropriate for telerehabilitation. It is also possible that a lack of experience or confidence providing telerehabilitation contributes to this perspective. Telerehabilitation has not historically been included in physical therapy curricula and most physical therapists had no experience providing telerehabilitation before the pandemic. Moreover, there is little to no evidence available in this area and best-practice recommendations for telerehabilitation are only now beginning to be published.^18^ Given the lack of telerehabilitation training, experience, evidence, and guidance, it is possible that physical therapists prefer providing care in-person because they are more familiar and comfortable with this approach.

Taken together, the survey results also suggest that patient preference plays an important role in determining how physical therapy care is provided. For example, a majority of physical therapists agreed that some patients may respond better to care provided by telerehabilitation and disagreed with a statement indicating that telerehabilitation should be used only when in-clinic care is not an option. About 80% of physical therapists indicated subsequently that telerehabilitation is more appropriate when patients have a preference for telerehabilitation. This is an important finding, as it suggests that there may not be a single answer to the question of whether telerehabilitation is an effective treatment for patients with musculoskeletal pain. Rather, the effectiveness of telerehabilitation as a care-delivery mechanism may be dependent on individual patient factors, such as patient preference. Researchers conducting studies examining the effectiveness of telerehabilitation might consider including subgroup analyses that examine the effects of telerehabilitation among those with preferences for or against telerehabilitation.

Strengths of this study are the use of a nationally distributed survey and access to a geographically diverse provider sample. The study also has important limitations to consider. As is common in studies using survey data, the results of our study may be influenced by recall bias, especially for questions that required survey respondents to recall information from prior to the pandemic. We also used a convenience sample of physical therapists belonging to the AOPT, which included a high percentage of board-certified physical therapists with 10 or more years of experience. It is possible that this sample is not representative of the full population of physical therapists caring for patients with musculoskeletal pain, which may limit the generalizability of our results. Our results may also have been influenced by response bias, as physical therapists with stronger opinions on telerehabilitation may have been more likely to respond to our survey invitation.

## Conclusions

Surveyed physical therapists reported providing care using telerehabilitation less frequently after the pandemic than during the pandemic, but at higher frequencies than prior to the pandemic. Physical therapists also reported that telerehabilitation can be an effective approach for treating patients with musculoskeletal pain, especially those who are unable to or prefer not to receive care in clinic. However, surveyed physical therapists do not view telerehabilitation as a replacement for in-clinic care. More research is needed to examine the effectiveness of care provided using telerehabilitation compared to in-clinic care to inform decisions made by physical therapists surrounding care delivery options.

## Supporting information

Suppemental Meterial

## Acknowledgment

For editorial assistance, we thank Denise Di Salvo, MS, in the Editorial Services group of The Johns Hopkins Department of Orthopaedic Surgery.

## Institutional Review Board Statement

This study was recognized as exempt by the Johns Hopkins Medicine Institutional Review Board (IRB00402769).

## Data Availability

Data included in this study are not publicly available but can be provided upon reasonable request to the corresponding author.

## References

1. Bierman RT, Kwong MW, Calouro C. State occupational and physical therapy telehealth laws and regulations: a 50-state survey. Int J Telerehabil. 2018;10(2):3–54. doi:10.5195/ijt.2018.6269

2. Lee AC, Davenport TE, Randall K. Telehealth physical therapy in musculoskeletal practice. J Orthop Sports Phys Ther. 2018;48(10):736–739. doi:10.2519/jospt.2018.0613

3. Trump Administration Issues Second Round of Sweeping Changes to Support U.S. Healthcare System During COVID-19 Pandemic | CMS. CMS Newsroom. April 30, 2020. Accessed October 3, 2024. https://www.cms.gov/newsroom/press-releases/trump-administration-issues-second-round-sweeping-changes-support-us-healthcare-system-during-covid

4. McLaughlin KH, Levy JF, Fritz JM, Skolasky RL. Trends in telerehabilitation utilization in the United States 2020-2021. Arch Phys Med Rehabil. 2024;105(7):1299–1304. doi:10.1016/j.apmr.2024.02.728

5. Seron P, Oliveros MJ, Gutierrez-Arias R, et al. Effectiveness of telerehabilitation in physical therapy: a rapid overview. Phys Ther. 2021;101(6):pzab053. doi:10.1093/ptj/pzab053

6. Suso-Martí L, La Touche R, Herranz-Gómez A, Angulo-Díaz-Parreño S, Paris-Alemany A, Cuenca-Martínez F. Effectiveness of Telerehabilitation in Physical Therapist Practice: An Umbrella and Mapping Review With Meta-Meta-Analysis. Phys Ther. 2021;101(5):pzab075. doi:10.1093/ptj/pzab075

7. Fritz JM, Lane E, Minick KI, et al. Perceptions of telehealth physical therapy among patients with chronic low back pain. Telemed Rep. 2021;2(1):258–263. doi:10.1089/tmr.2021.0028

8. Fritz JM, Minick KI, Brennan GP, et al. Outcomes of telehealth physical therapy provided using real-time, videoconferencing for patients with chronic low back pain: a longitudinal observational study. Arch Phys Med Rehabil. 2022;103(10):1924–1934. doi:10.1016/j.apmr.2022.04.016

9. Skolasky RL, Kimball ER, Galyean P, et al. Identifying perceptions, experiences, and recommendations of telehealth physical therapy for patients with chronic low back pain: a mixed methods survey. Arch Phys Med Rehabil. 2022;103(10):1935–1943. doi:10.1016/j.apmr.2022.06.006

10. Malliaras P, Merolli M, Williams CM, Caneiro JP, Haines T, Barton C. ‘It’s not hands-on therapy, so it’s very limited’: Telehealth use and views among allied health clinicians during the coronavirus pandemic. Musculoskelet Sci Pract. 2021;52:102340. doi:10.1016/j.msksp.2021.102340

11. Haines KJ, Sawyer A, McKinnon C, et al. Barriers and enablers to telehealth use by physiotherapists during the COVID-19 pandemic. Physiotherapy. 2023;118:12–19. doi:10.1016/j.physio.2022.09.003

12. Carter SK, Rizzo JA. Use of outpatient physical therapy services by people with musculoskeletal conditions. Phys Ther. 2007;87(5):497–512. doi:10.2522/ptj.20050218

13. Freburger JK, Holmes GM, Carey TS. Physician referrals to physical therapy for the treatment of musculoskeletal conditions. Arch Phys Med Rehabil. 2003;84(12):1839–1849. doi:10.1016/s0003-9993(03)00375-7

14. Burns SA, Cleland JA, Rivett DA, Snodgrass SJ. Examination procedures and interventions for the hip in the management of low back pain: a survey of physical therapists. Braz J Phys Ther. 2019;23(5):419–427. doi:10.1016/j.bjpt.2018.09.007

15. McLaughlin KH, Archer KR, Shafiq B, Wegener ST, Reider L. Orthopedic surgeons and physical therapists differ regarding rehabilitative needs after lower extremity fracture repair. Physiother Theory Pract. 2023;39(11):2446–2453. doi:10.1080/09593985.2022.2078753

16. Magel J, Bishop MD, Lonnemann E, et al. The association between advanced orthopedic certification and confidence and engagement in prescription opioid medication misuse management practices: a cross-sectional study. J Man Manip Ther. 2022;30(4):228–238. doi:10.1080/10669817.2021.2000818

17. Werneke MW, Deutscher D, Grigsby D, Tucker CA, Mioduski JE, Hayes D. Telerehabilitation during the COVID-19 pandemic in outpatient rehabilitation settings: a descriptive study. Phys Ther. 2021;101(7):pzab110. doi:10.1093/ptj/pzab110

18. Lee AC, Deutsch JE, Holdsworth L, et al. Telerehabilitation in physical therapist practice: a clinical practice guideline from the American Physical Therapy Association. Phys Ther. 2024;104(5):pzae045. doi:10.1093/ptj/pzae045

