## Supplementary material for "Physical therapists’ perceptions of telerehabilitation for patients with musculoskeletal conditions in a post-pandemic world": Suppemental Meterial

**Physical Therapist Perceptions of Telerehabilitation Post-Pandemic: A National Provider Survey**

The COVID-19 pandemic facilitated rapid expansion of telehealth services in the United States. However, with the recent end of the Public Health Emergency, it is unclear how telehealth will and should be used moving forward. This survey is being conducted to examine physical therapists’ attitudes and beliefs surrounding care delivered by telehealth, also referred to as *telerehabilitation*. This survey takes approximately 6 minutes to complete.

Completion of this survey will serve as your consent to participate in this study. All responses will be collected anonymously. Please note that this survey has been mailed to members of the Academy of Orthopaedic Physical Therapists and members of the American Academy of Orthopaedic Manual Physical Therapists. If you belong to both of these groups, please only complete this survey once.

Please note that in this survey, the term *telerehabilitation* is used to refer to physical therapy care delivered remotely, using synchronous video or audio-only visits.

***Section 1: Telerehabilitation Experience Before, During and After the COVID-19 Pandemic***

*For each question, please select the response that most closely describes your previous or current use of telerehabilitation.*

1. Prior to the beginning of the COVID-19 pandemic in March 2020, how often did you provide care via telerehabilitation?
   a. Never
   b. Rarely (Less than once a month)
   c. Sometimes (At least once a month)
   d. Often (At least weekly)
   e. Very frequently (3 or more days per week)
2. Prior to the COVID-19 pandemic, what proportion of patient visits did you complete via telerehabilitation during a typical week?
   a. 0%
   b. 1-10%
   c. 10-25%
   d. 25-50%
   e. >50%
3. Which of the following best describes the highest frequency with which you used telerehabilitation during the COVID-19 pandemic (March 2020-May 2023)?
   a. Never
   b. Rarely (Less than once a month)
   c. Sometimes (At least once a month)
   d. Often (At least weekly)
   e. Very frequently (3 or more days per week)
4. Which of the following approximates the highest proportion of patients visits you completed via telerehabilitation in a week during the COVID-19 pandemic (March 2020-May 2023)?
   a. 0%
   b. 1-10%
   c. 10-25%
   d. 25-50%
   e. >50%
5. Since the Public Health Emergency ended in May 2023, how often do you typically see any patients via telerehabilitation?
   a. Never
   b. Rarely (Less than once a month)
   c. Sometimes (At least once a month)
   d. Often (At least weekly)
   e. Very frequently (3 or more days per week)
6. Since Public Health Emergency ended in May 2023, what proportion of patients do you see via telerehabilitation during a typical week?
   a. 0%
   b. 1-10%
   c. 10-25%
   d. 25-50%
   e. >50%

***Section 2: Perceptions of Telerehabilitation***

*Please indicate your level of agreement with each of the following statements.*

7. I can deliver effective care for patients with musculoskeletal pain through telerehabilitation.
   a. Strongly agree (5)
   b. Agree (4)
   c. Neutral (3)
   d. Disagree (2)
   e. Strongly disagree (1)
8. I can deliver care through telerehabilitation that is equally as effective as in-clinic care for most of the patients I see with musculoskeletal pain.
   a. Strongly agree (5)
   b. Agree (4)
   c. Neutral (3)
   d. Disagree (2)
   e. Strongly disagree (1)
9. I believe certain patients may respond better to care I provide through telerehabilitation versus in-clinic care.
   a. Strongly agree (5)
   b. Agree (4)
   c. Neutral (3)
   d. Disagree (2)
   e. Strongly disagree (1)
10. I would feel comfortable providing a full episode of care through telerehabilitation if that was my patient’s preference
    a. Strongly agree (5)
    b. Agree (4)
    c. Neutral (3)
    d. Disagree (2)
    e. Strongly disagree (1)
11. I would prefer to provide telerehabilitation as part of a hybrid model (combination of in-clinic and virtual care) for patients that express a desire to utilize telerehabilitation, rather than exclusively through telerehabilitation
    a. Strongly agree (5)
    b. Agree (4)
    c. Neutral (3)
    d. Disagree (2)
    e. Strongly disagree (1)
12. Telerehabilitation should only be utilized when in-clinic care is not an option.
    a. Strongly agree (5)
    b. Agree (4)
    c. Neutral (3)
    d. Disagree (2)
    e. Strongly disagree (1)
13. Telerehabilitation is not an appropriate way to deliver care for patients, regardless of their ability to attend in-clinic visits.
    a. Strongly agree (5)
    b. Agree (4)
    c. Neutral (3)
    d. Disagree (2)
    e. Strongly disagree (1)

***Section 3: Future Telerehabilitation Use***

*For the following questions, please assume that telerehabilitation will continue to be reimbursed at equivalent rates to in-clinic physical therapy.*

*Please indicate your level of agreement with each of the following statements.*

14. I plan to provide telerehabilitation for patients with musculoskeletal pain in the future.
    a. Strongly agree (5)
    b. Agree (4)
    c. Neutral (3)
    d. Disagree (2)
    e. Strongly disagree (1)
15. Telerehabilitation plays an important role in expanding access to physical therapy in the United States.
    a. Strongly agree (5)
    b. Agree (4)
    c. Neutral (3)
    d. Disagree (2)
    e. Strongly disagree (1)
16. Please indicate to what extent each of the following items is a barrier to using telerehabilitation among patients with musculoskeletal pain at your clinic/hospital.

|  | **Not a barrier** | **Somewhat of a barrier** | **Moderate barrier** | **Extreme barrier** |
| --- | --- | --- | --- | --- |
| Patient internet access |  |  |  |  |
| Patients experience issues with telehealth technology |  |  |  |  |
| You (provider) experience issues with telehealth technology |  |  |  |  |
| Lack of training on how to provide care via telehealth |  |  |  |  |

***Section 4. Patient Subgroups Appropriate for Telerehabilitation***

*Please indicate your level of agreement with the following statement.*

17. Telerehabilitation is best suited for specific patient groups of patients with musculoskeletal pain
    a. Strongly agree (5)
    b. Agree (4)
    c. Neutral (3)
    d. Disagree (2)
    e. Strongly disagree (1)
18. Please indicate how you believe each of the following scenarios or details would influence the appropriateness of a patient for telerehabilitation

| ***The patient…*** | **More appropriate for telerehabilitation (3)** | **Less appropriate for telerehabilitation (2)** | **No impact on appropriateness (1)** |
| --- | --- | --- | --- |
| is available to attend in-clinic visits |  |  |  |
| prefers televisits |  |  |  |
| has access to the internet |  |  |  |
| has a chronic condition |  |  |  |
| has an acute condition |  |  |  |
| has high psychosocial risk |  |  |  |
| has low psychosocial risk |  |  |  |
| has high self-efficacy |  |  |  |
| has low self-efficacy |  |  |  |
| has extremity pain |  |  |  |
| has spine pain |  |  |  |
| is post-operative |  |  |  |
| is comfortable using technology |  |  |  |
| is relatively young |  |  |  |
| is relatively older |  |  |  |

19. Please enter any patient factors not included in the previous question that you believe should be considered when determining the appropriateness of telerehabilitation for a specific patient: [FREE TEXT]

***Section 5: Respondent Characteristics***

20. What setting do you primarily work in?
    a. Acute care hospital
    b. Health-system or hospital-based outpatient clinic
    c. Private outpatient practice or group practice
    d. Skilled nursing facility/extended or intermediate care facility
    e. School system (preschool, primary, secondary)
    f. Academic institution (post-secondary)
    g. Health and wellness facility
    h. Research center
    i. Industry
21. Which of the following best describes the region you practice in?
    a. a. South Atlantic (DE, MD, DC, GA, NC, PR, SC, VA, WV, FL)
    b. Middle Atlantic (NJ, NY, PA)
    c. East North Central (IL, MI, OH, WI)
    d. West North Central (IA, KS, MN, MO, NE, ND, SD)
    e. East South Central (AL, KY, MS, TN)
    f. West South Central (AR, LA, OK, TX)
    g. New England (CT, ME, MA, NH, RI, VT)
    h. Pacific (AK, CA, HI, OR, WA)
    i. Mountain (AZ, CO, ID, MT, NV, NM, UT, WY)
22. Which of the following best describes the area that you practice in?
    a. Urban
    b. Suburban
    c. Rural

23. What percentage of your time do you spend on direct patient care?
    a. 0-25%
    b. 25-50%
    c. 50-75%
    d. >75%
24. What percentage of your clinical caseload is comprised of patients with musculoskeletal pain?
    a. 0-25%
    b. 25-50%
    c. 50-75%
    d. >75%
25. How do you describe yourself?
    a. Female
    b. Male
    c. Non-binary/third gender
    d. Prefer to self-describe: ______________________
    e. Prefer not to say
26. How many years have you been a practicing physical therapist?
    a. < 5 years
    b. 5-10 years
    c. 10-20 years
    d. > 20 years
27. What is the highest physical therapy-related degree you have earned?
    a. Certificate
    b. Bachelors
    c. Masters
    d. DPT/tDPT/DScPT
    e. PhD, ScD
28. Are you certified as an American Board of Physical Therapy Specialist (ABPTS)?
    a. Yes
    b. No

If yes, which specialization?

a. Orthopaedics
b. Sports
c. Faculty
d. Pediatrics
e. Women’s Health
f. Neurology
g. Cardiopulmonary
h. Acute Care
i. Wound Care
29. Are you fellowship trained or currently enrolled in a fellowship program?
    a. Yes
    b. No

If yes, which of the following?

a. Orthopaedic manual physical therapy
b. Sports
c. Spine
d. Education leadership
e. Movement science
f. Upper extremity athlete
g. Critical care
h. Hand therapy
i. Neonatology
